# Direction of apoB/LDL-C Discordance and Systemic Inflammation in U.S. Adults

**DOI:** 10.64898/2026.11.12.26367975

**Authors:** Ose Kadiri, Iretomiwa Esho

**Affiliations:** Department of Internal Medicine, St. Elizabeth Youngstown Hospital, Youngstown, OH; The MITRE Corporation, McLean, VA

**Keywords:** apolipoprotein B, LDL cholesterol, discordance, inflammation

## Abstract

**Background:** Apolipoprotein B (apoB) and low-density lipoprotein cholesterol (LDL-C) are established biomarkers of atherosclerotic cardiovascular disease (ASCVD) that tend to be closely correlated. However, some individuals have discordant apoB and LDL-C. We evaluated whether apoB/LDL-C discordance is associated with elevated high-sensitivity C-reactive protein (hs-CRP), a key marker of systemic inflammation, in a nationally representative sample.

**Methods and Results:** We analyzed cross-sectional data of 2226 adults aged 20 years or older from the 2015 to 2016 National Health and Nutrition Examination Survey with complete apoB, LDL-C, and high-sensitivity C-reactive protein (hs-CRP). LDL-C and apoB were converted to survey-weighted *z* scores, and discordance was the residual of apoB regressed on LDL-C. Participants were classified as apoB-high, LDL-C-high, or concordant (reference). The association of discordance group with elevated hs-CRP (*>*3 mg/L) was estimated with survey-weighted logistic regression, adjusted for age, sex, body mass index (BMI), and diabetes. Discordance was present in 26.4% of adults (13.7% apoB-higher, 12.6% LDL-C-higher). Elevated hs-CRP prevalence differed by discordance group: 47.6% (95% CI, 37.6–57.7) in the apoB-high group versus 20.5% (12.9–28.1) in the LDL-C-high group and 33.6% (29.7–37.4) in the concordant group. Relative to concordant participants, adjusted odds of elevated hs-CRP were higher for apoB-high discordance (odds ratio [OR], 1.58; 95% CI, 1.03–2.45) and lower for LDL-C-high discordance (OR, 0.49; 95% CI, 0.30–0.78). Similar results were obtained among participants not on statin therapy, with apoB-high discordance demonstrating stronger adjusted odds of elevated hs-CRP (OR, 2.02; 95% CI, 1.01–4.04).

**Conclusion:** Discordantly high apoB relative to LDL-C is associated with systemic inflammation in a nationally representative sample of U.S. adults.

## Introduction

Lowering LDL cholesterol (LDL-C) is central to the prevention of atherosclerotic cardiovascular disease (ASCVD), and LDL-C remains the principal target of lipid-lowering guidelines world-wide [1–3]. The causal role of LDL and other apolipoprotein B (apoB)-containing lipoproteins in atherogenesis is supported by genetic, epidemiologic, and clinical trial evidence [4]. However, LDL-C quantifies the cholesterol mass carried within circulating lipoproteins, not the number of atherogenic particles. Because each atherogenic lipoprotein, including very-low-density lipoprotein, intermediate-density lipoprotein, LDL, and lipoprotein(a), carries a single apoB molecule, apoB provides a direct count of the atherogenic particle burden [5, 6].

When cholesterol mass and particle number diverge, LDL-C and apoB become discordant [5]. Multiple cohorts have shown that when apoB and LDL-C disagree, cardiovascular risk tracks with apoB rather than with LDL-C, and that discordant elevation of apoB predicts incident events and subclinical atherosclerosis beyond LDL-C [5, 7–9]. These observations have contributed to the in-corporation of apoB into clinical guidelines as a risk-enhancing factor and an alternative measure of atherogenic burden [2, 3]. Recent nationally representative data indicate that apoB/LDL-C discordance is common in the general population [10].

In parallel, systemic inflammation has emerged as an independent and modifiable contributor to ASCVD [11]. High-sensitivity C-reactive protein (hs-CRP) is a validated marker of inflammatory risk that predicts cardiovascular events across the spectrum of LDL-C [12–14], and an hs-CRP concentration greater than 3 mg/L identifies individuals at higher vascular risk [12]. Anti-inflammatory therapy reduces cardiovascular events independent of lipid lowering [15–17], and among statin-treated patients, inflammation assessed by hs-CRP predicts residual risk at least as strongly as LDL-C [18]. A 2025 American College of Cardiology Scientific Statement summarizes the growing evidence base linking inflammation to cardiovascular disease and its assessment in clinical care [19].

The metabolic profile that produces an elevation of apoB relative to LDL-C may also promote inflammation. Remnant and triglyceride-rich lipoproteins have been linked to causing cause low-grade inflammation [20, 21], so it is plausible that the direction of apoB/LDL-C discordance (more specifically, apoB-high discordance) carries information about concurrent inflammation. Previous discordance research has focused on atherosclerotic events and subclinical disease rather than inflammatory burden, and it has not been examined whether discordance is associated with low-grade inflammation in the general population.

We used the National Health and Nutrition Examination Survey (NHANES) to determine whether the direction of apoB/LDL-C discordance is associated with low-grade systemic inflammation among U.S. adults, and to examine the extent to which adiposity and sex account for that association.

## Methods

### Data availability

NHANES data are publicly available from the National Center for Health Statistics [22]. Analytic code will be made available upon reasonable request.

### Data source and study population

We analyzed the 2015-2016 cycle of NHANES, a continuous, cross-sectional survey of the civilian noninstitutionalized US population that uses a complex, multistage probability sampling design. Details of recruitment, examination, and laboratory procedures are described elsewhere [23]. Note that the 2015-2016 cycle is the only NHANES cycle with concurrent measurements of both apoB and hs-CRP.

The analytic sample comprised adults aged 20 years or older with complete data for apoB, LDL-C, hs-CRP, and survey design variables. Because apoB and calculated LDL-C were measured in the morning fasting subsample, eligibility required participation in that subsample. The participant flowchart is shown in Figure 1. The final analytic sample included 2,226 participants. Adjusted models were restricted to the 2,151 participants with complete covariate data.

**Figure 1.**
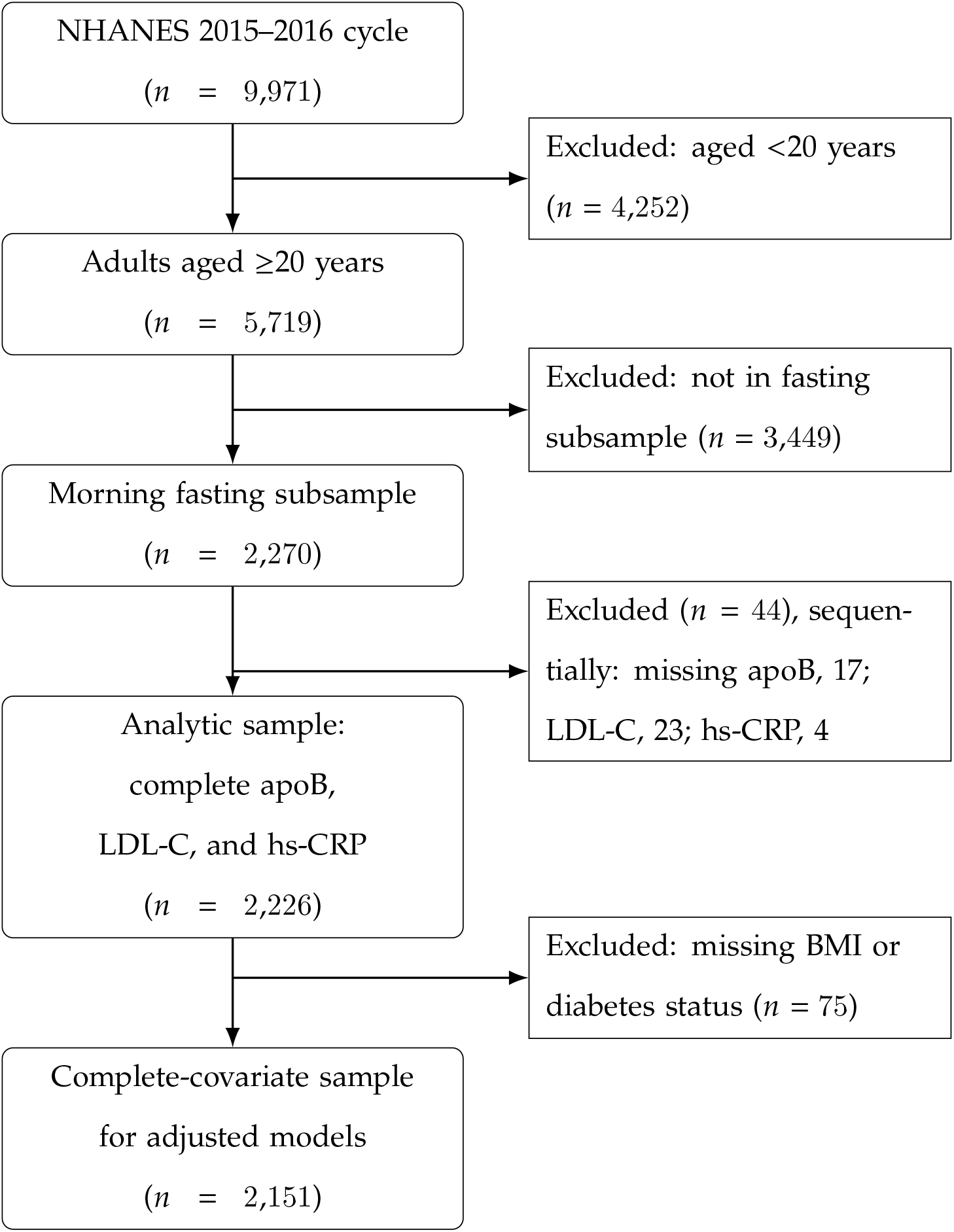
Derivation of the analytic sample from the NHANES 2015 to 2016 cycle. Counts are unweighted participants.

### Laboratory measures

apoB was measured by immunoturbidimetric assay and hs-CRP by particle immunoassay on automated analyzers. LDL-C was the NHANES-provided value calculated using the Friedewald equation. Full laboratory methods are available in the NHANES documentation [22].

### Definition of discordance

There is no universally accepted definition of discordance in the literature. Existing approaches fall into three broad classes: cut points [10, 24], population percentiles [25], and regression residuals [26, 27]. We adopt the residual-based approach here: LDL-C and apoB were each converted to survey-weighted *z* scores using the weighted mean and standard deviation, and the apoB *z* score was regressed on the LDL-C *z* score. Because both variables were standardized, the slope equals the weighted correlation (*r*), and the expected apoB *z* score for a given LDL-C *z* score is *r* times the LDL-C *z* score. The residual (observed minus expected apoB *z* score) was divided by its expected standard deviation, 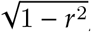, to yield a residual discordance *z* score, *€*_*z*_, on an interpretable scale.

Participants were classified using an absolute residual discordance *z* score threshold of 1:

- **apoB higher than LDL-C**: *€*_*z*_ ≥ +1
- **LDL-C higher than apoB**: *€*_*z*_ ≤ −1
- **Concordant** (reference): |*€*_*z*_| *<* 1

This method asks whether a participant’s apoB is higher or lower than expected given the participant’s LDL-C.

### Outcome

The outcome was elevated hs-CRP, defined as an hs-CRP concentration greater than 3 mg/L, consistent with the American Heart Association and Centers for Disease Control and Prevention threshold for higher relative vascular risk [12].

### Statistical analysis

Analyses were conducted in R using the survey package. Because apoB and calculated LDL-C are measured in the morning fasting subsample, all estimates used the NHANES fasting subsample 2-year sampling weights (WTSAF2YR).

The primary analysis was a logistic regression model with elevated hs-CRP as the dependent variable and discordance group as the independent variable, entered as two indicator terms with the concordant group as the reference. We report an unadjusted model and a model adjusted for age, sex, BMI, and diagnosed diabetes.

To identify which covariates accounted for the change in the adjusted estimates, we fit a sequence of nested models on a single fixed sample of participants with complete covariate data, adding covariates one at a time. Holding the sample constant ensures that any change in the odds ratio reflects the covariate added rather than a change in sample composition. A two-sided *P* value below 0.05 was considered statistically significant. In a prespecified sensitivity analysis, we repeated the unadjusted and adjusted models after excluding participants taking a statin (atorvastatin, simvastatin, rosuvastatin, pravastatin, lovastatin, fluvastatin, or pitavastatin).

## Results

### Sample characteristics

The mean LDL-C was 112.6 (SD 35.7) mg/dL and mean apoB was 93.2 (SD 25.5) mg/dL. A scatter plot of the weighted apoB z score versus LDL-C z score is shown in Fig. 2. As expected, LDL-C and apoB were strongly correlated (*r* = 0.924; *R*^2^ = 0.853). The discordant groups, representing participants with |*€*_*z*_| ≥ 1 are depicted above and below the main cloud that represents the concordant group. The overall survey-weighted prevalence of elevated hs-CRP was 33.8% (95% CI, 29.8–37.9). Survey-weighted characteristics by discordance group are shown in Table 1.

**Table 1:** Weighted characteristics of US adults by apoB/LDL-C discordance group.

| Characteristic | Overall | Concordant | apoB higher | LDL-C higher |
|---|---|---|---|---|
| Unweighted, *n* | 2,151 | 1,597 | 324 | 230 |
| Age, y | 47.4 | 47.3 | 53.2 | 42.2 |
| Male, % | 47.9 | 48.0 | 56.7 | 37.4 |
| BMI, kg/m<sup>2</sup> | 29.4 | 29.3 | 31.5 | 28.0 |
| Diagnosed diabetes, % | 10.9 | 8.8 | 30.0 | 1.9 |
| Statin use, % | 18.6 | 16.9 | 41.1 | 4.5 |
| Elevated hs-CRP (>3 mg/L), % | 33.3 | 33.1 | 47.4 | 19.4 |
| LDL-C, mg/dL | 112.6 | 110.4 | 117.5 | 120.1 |
| apoB, mg/dL | 93.1 | 90.7 | 113.7 | 84.7 |

Analytic sample with complete covariate data (*n* = 2,151). Values are survey-weighted means or percentages. BMI indicates body mass index; hs-CRP, high-sensitivity C-reactive protein; LDL-C, low-density lipoprotein cholesterol. Elevated hs-CRP percentages are computed on the complete-covariate sample and differ marginally from Table 2 (*n* = 2,226).

**Figure 2.**
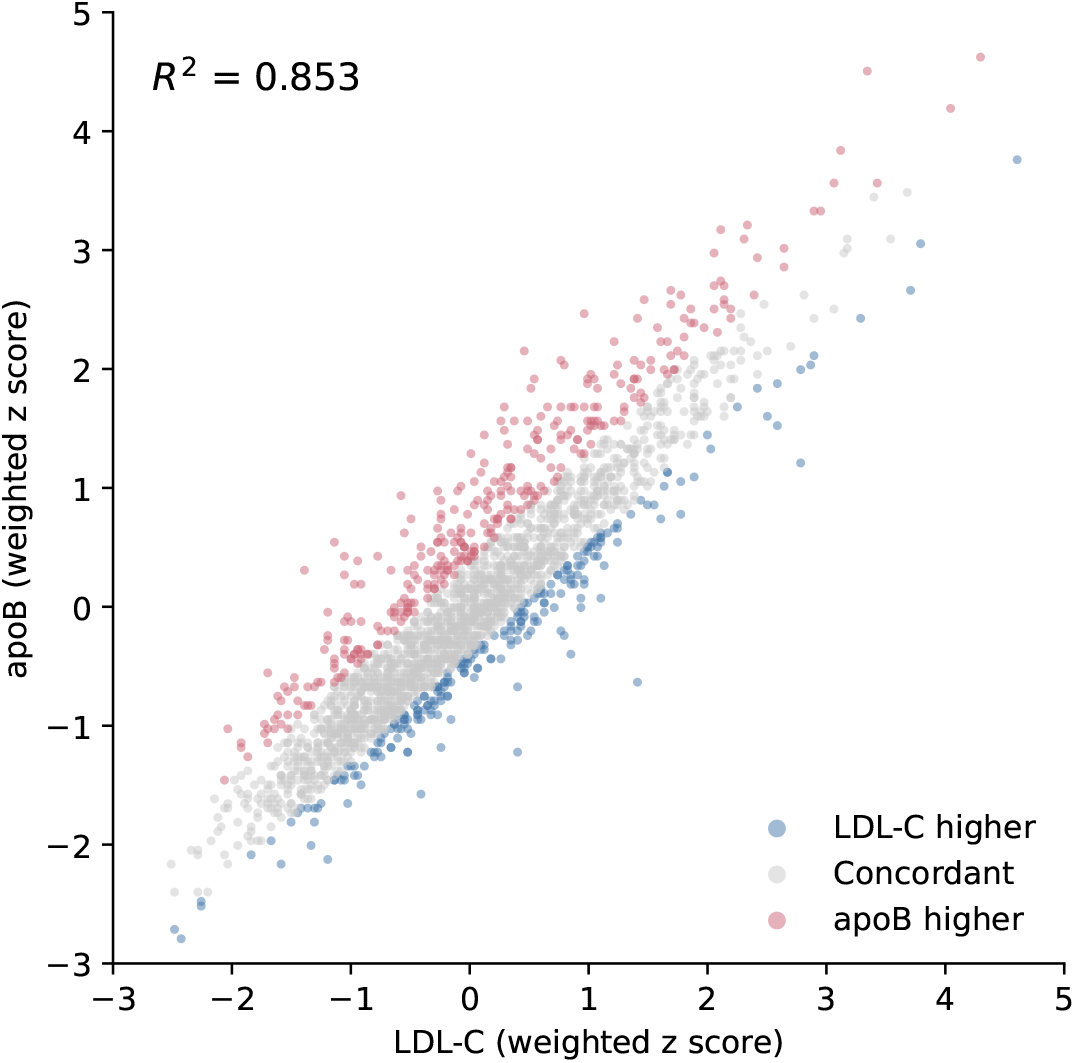
Weighted apoB *z* score versus LDL-C *z* score in US adults (*n* = 2,226; *R*^2^ = 0.853), with participants colored by discordance group. Points with apoB higher than their LDL-C predicts (apoB higher) sit above the main cloud and those with lower apoB (LDL-C higher) sit below.

The three discordance groups differed in cardiometabolic profile. Compared with the concordant group, the apoB-high group had higher BMI (31.5 versus 29.3 kg/m^2^), a greater proportion of diagnosed diabetes (30.0% versus 8.8%), older mean age (53.2 versus 47.3 years), and a higher proportion of men (56.7% versus 48.0%). The LDL-C-high group had the most favorable profile on these measures, including the lowest BMI (28.0 kg/m^2^), lowest diabetes prevalence (1.9%), youngest mean age (42.2 years), and lowest proportion of men (37.4%).

apoB/LDL-C discordance was present in 26.4% of adults at the absolute residual *z* score threshold of 1, with the two directions of similar frequency: 13.7% had apoB higher than LDL-C and 12.6% had LDL-C higher than apoB; the remaining 73.6% were concordant (Table 2).

**Table 2:** Weighted prevalence of discordance groups and of elevated hs-CRP within groups.

| Group | Unweighted, *n* | Prevalence of group, % | Elevated hs-CRP, % (95% CI) |
|---|---|---|---|
| Concordant (reference) | 1,652 | 73.6 | 33.6 (29.7–37.4) |
| apoB higher than LDL-C | 341 | 13.7 | 47.6 (37.6–57.7) |
| LDL-C higher than apoB | 233 | 12.6 | 20.5 (12.9–28.1) |

Full analytic sample (*n* = 2,226). hs-CRP indicates high-sensitivity C-reactive protein.

### Elevated hs-CRP by discordance group

The prevalence of elevated hs-CRP differed substantially and directionally across groups (Table 2; Figure 3). It was highest in the apoB-high group at 47.6% (95% CI, 37.6–57.7), intermediate in the concordant group at 33.6% (29.7–37.4), and lowest in the LDL-C-high group at 20.5% (12.9–28.1).

**Figure 3.**
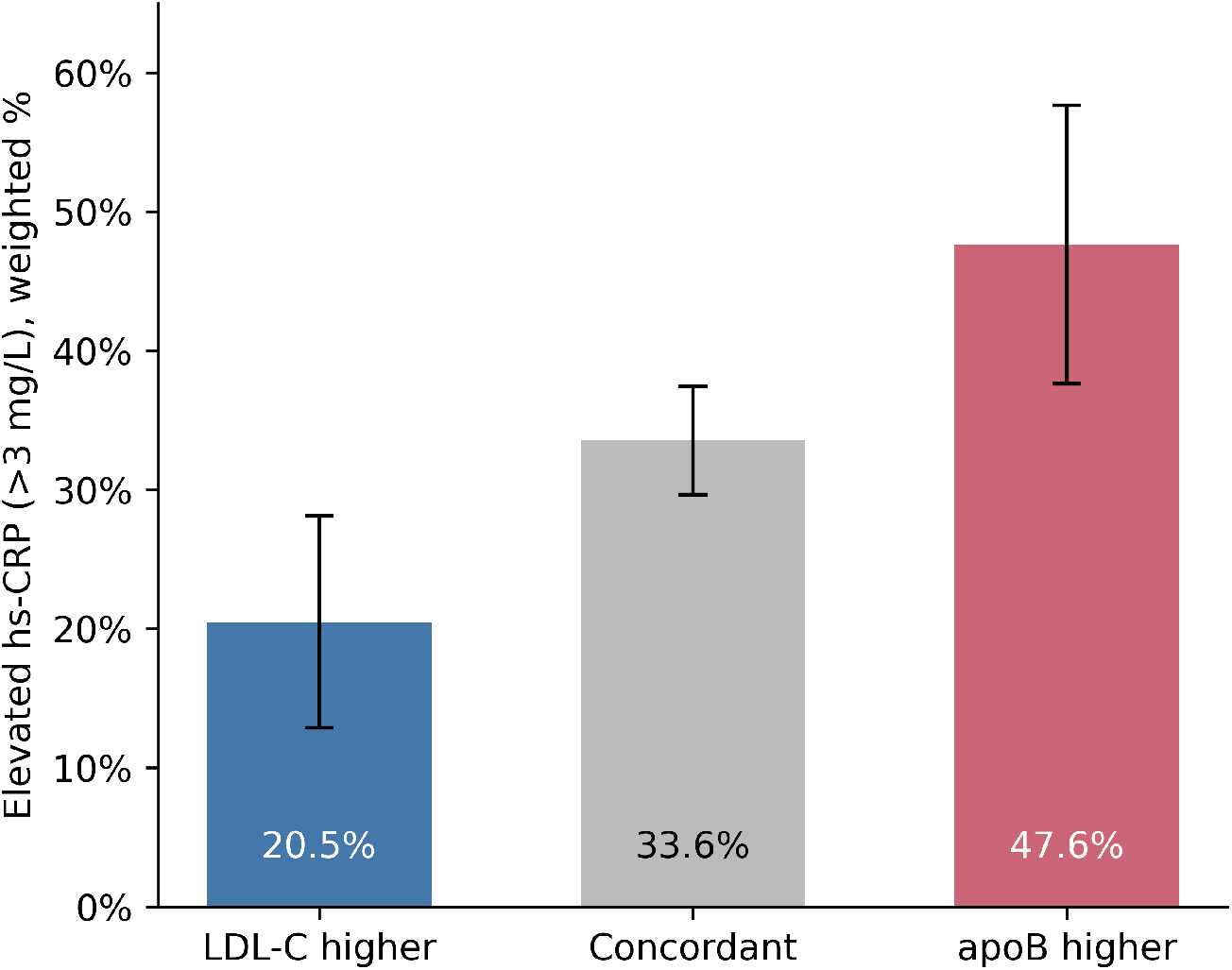
Survey-weighted prevalence of elevated high-sensitivity C-reactive protein (*>*3 mg/L) by apoB/LDL-C discordance group, with 95% CIs.

### Association between directional discordance and elevated hs-CRP

In the unadjusted model, the odds of elevated hs-CRP were higher for apoB-high discordance (OR, 1.80; 95% CI, 1.21–2.67; *P* = 0.006) and lower for LDL-C-higher discordance (OR, 0.51; 95% CI, 0.32–0.81; *P* = 0.007) (Table 3) relative to concordant participants.

**Table 3:** Survey-weighted logistic regression for elevated hs-CRP (*>*3 mg/L).

| Term | Unadjusted OR (95% CI) | *P* | Adjusted OR (95% CI) | *P* |
|---|---|---|---|---|
| apoB higher than LDL-C | 1.80 (1.21–2.67) | 0.006 | 1.58 (1.03–2.45) | 0.039 |
| LDL-C higher than apoB | 0.51 (0.32–0.81) | 0.007 | 0.49 (0.30–0.78) | 0.006 |

Concordant group is the reference. Unadjusted model *n* = 2,226; adjusted model *n* = 2,151, additionally adjusted for age, sex, BMI, and diagnosed diabetes (estimates not shown). BMI indicates body mass index; OR, odds ratio.

After adjustment for age, sex, BMI, and diagnosed diabetes (*n* = 2,151), the odds of elevated hs-CRP remained higher for apoB-high discordance (OR, 1.58; 95% CI, 1.03–2.45; *P* = 0.039) and lower for LDL-C-higher discordance (OR, 0.49; 95% CI, 0.30–0.78; *P* = 0.006) (Table 3).

### Contributions of adiposity and sex to the adjusted estimate

Using a stepwise adjustment on a fixed sample, we found that the odds ratio for the apoB-high group fluctuated across models (Table 4). Starting from the crude estimate (*OR* = 1.82), adding BMI dropped the odds ratio to 1.50, whereas further adding sex raised it back to 1.62. Age and diabetes had minimal impact. We can understand these opposing shifts by examining the participant profiles. Participants in the apoB-high group had a higher average BMI than the reference group (31.5 vs. 29.3 kg/m^2^). Because higher BMI correlates with elevated hs-CRP, adjusting for BMI removed positive confounding and lowered the odds ratio. Conversely, the apoB-high group had a larger proportion of men (56.7% vs. 48.0%), and men were much less likely to have elevated hs-CRP than women (27.2% vs. 38.9%). This surplus of lower-CRP men artificially attenuated the crude odds ratio, and adjusting for sex removed this artifact.

**Table 4:** Sequential adjustment of the discordance-inflammation association.

| Model | apoB higher, OR | *P* | LDL-C higher, OR | *P* |
|---|---|---|---|---|
| Crude | 1.82 | 0.006 | 0.49 | 0.005 |
| + BMI | 1.50 | 0.084 | 0.53 | 0.015 |
| + BMI + age | 1.47 | 0.082 | 0.53 | 0.018 |
| + BMI + diabetes | 1.48 | 0.087 | 0.53 | 0.016 |
| + BMI + sex | 1.62 | 0.043 | 0.48 | 0.005 |
| + BMI + sex + age + diabetes | 1.58 | 0.039 | 0.49 | 0.006 |

All models fit on the fixed complete-covariate sample (*n* = 2,151) so estimates are directly comparable across rows.
BMI indicates body mass index; OR, odds ratio.

### Sensitivity to statin therapy

Since statin therapy can significantly reduce both LDL-C and hs-CRP, we conducted a sensitivity analysis to exclude participants on statins. As seen in Table 1, statin use differed significantly across groups. 41.1% of the apoB-higher group were taking a statin, compared with 16.9% of the concordant group and 4.5% of the LDL-C-higher group. This gradient is expected because statins lower LDL-C proportionally more than apoB [5, 28] and therefore shift statin-treated patients toward discordantly high apoB. Because statins also lower hs-CRP, their concentration in the apoB-high group could further obscure the observed association.

To address the effect of statins, we repeated the analysis among statin-naive participants (*n* = 1,753; adjusted model *n* = 1,711). Notably, the pattern persisted and even strengthened in the apoB-higher group. Compared to concordant participants, those with apoB-higher discordance had higher odds of elevated hs-CRP (crude OR, 2.38; 95% CI, 1.41–4.00; adjusted OR, 2.02; 95% CI, 1.01–4.04) and lower for LDL-C-higher discordance (adjusted OR, 0.48; 95% CI, 0.28–0.81). This stronger association aligns with our expectation. Because statin users cluster in the apoB-high group and have pharmacologically lowered hs-CRP, including them in the full sample biases the apoB-higher association toward the null. Excluding participants on statins removes this masking effect and offers a cleaner, though less precise (due to reduced sample size) estimate. These results are summarized in Table 5.

**Table 5:** Association of discordance group with elevated hs-CRP (*>*3 mg/L), in the full sample and among statin-naive participants.

| Discordance group | Full sample |  | Statin-naive |  |
|---|---|---|---|---|
|  | Unadjusted<br>OR (95% CI) | Adjusted<br>OR (95% CI) | Unadjusted<br>OR (95% CI) | Adjusted<br>OR (95% CI) |
| apoB higher than LDL-C | 1.80 (1.21–2.67) | 1.58 (1.03–2.45) | 2.38 (1.41–4.00) | 2.02 (1.01–4.04) |
| LDL-C higher than apoB | 0.51 (0.32–0.81) | 0.49 (0.30–0.78) | 0.49 (0.31–0.79) | 0.48 (0.28–0.81) |

Concordant group is the reference. Full-sample unadjusted *n* = 2,226, adjusted *n* = 2,151; statin-naive unadjusted *n* = 1,753, adjusted *n* = 1,711. Adjusted models include age, sex, BMI, and diagnosed diabetes (estimates not shown). Statin users were identified from the prescription-medication file. BMI indicates body mass index; hs-CRP, high-sensitivity C-reactive protein; OR, odds ratio.

## Discussion

apoB elevation relative to LDL-C reflects an excess of cholesterol-depleted particles such as small dense LDL and remnant lipoproteins [5, 7]. Elevated remnant cholesterol has been linked to low-grade inflammation, [20, 21], and it is plausible that individuals with discordantly high apoB carry more of these inflammation-promoting lipoproteins. Our results are consistent with this account by showing that individuals with discordantly high apoB also exhibited greater inflammatory burden. Prior discordance studies have primarily related apoB elevation to atherosclerotic events and subclinical atherosclerosis [7–9, 29]. By using elevated hs-CRP as the outcome, we show that high apoB relative to LDL-C identifies individuals with a substantially higher burden of systemic inflammation, whereas discordantly high LDL-C relative to apoB identifies individuals with a lower burden of chronic inflammation. This suggests concurrent low-grade inflammation as a candidate mechanism accompanying the excess risk previously attributed to discordant apoB.

An observation of our analysis is the opposing influence of adiposity and sex. BMI functioned as a positive confounder. Because the apoB-high group had a higher mean BMI and adiposity drives hs-CRP, adjusting for BMI attenuated the association. Conversely, sex acted as a suppressor variable. The apoB-higher group was predominantly male, and since men tend to have lower hs-CRP than women [30, 31], controlling for sex strengthened the association between apoB-high discordance and elevated hs-CRP.

We consider the effect of statin therapy on our analysis. In Table 1, we showed that statin use differed significantly across the groups, from 4.5% in the LDL-C-high group to 41.1% in the apoB-higher group. This pattern is expected because statins lower LDL-C more effectively than apoB [5, 28], ultimately producing a relative excess of individuals with elevated apoB relative to LDL-C. As a result of this concentration of statin users in the apoB-high group (who also are expected to have reduced hs-CRP from statin therapy), the association of apoB-high discordance with elevated inflammation was weakened. Therefore, the full-sample estimate is biased toward the null, and the association is in fact larger among statin-naive participants (adjusted OR, 2.02 versus 1.58).

Our findings may have clinical relevance. In patients whose apoB exceeds expectations set by LDL-C, residual cholesterol risk and inflammatory risk may coexist, thus identifying individuals for whom both particle-directed and inflammation-directed strategies could be considered.

Anti-inflammatory therapies reduce cardiovascular events [15–17], and inflammation assessed by hs-CRP predicts residual risk in treated patients [18], so recognizing an inflammation-associated lipoprotein phenotype could inform risk stratification. Lastly, by showing that elevated hs-CRP aligns with discordantly high apoB, our findings support existing literature indicating that apoB captures cardiovascular risk more completely than LDL-C [7].

### Limitations

This study has a few limitations. First, the design is cross-sectional and based on a single NHANES cycle, so we cannot infer whether discordantly high apoB causes low-grade inflammation or vice versa. Secondly, estimates produced here may not generalize to other periods or populations. Furthermore, hs-CRP was measured once, and a single measurement is subject to variation. Finally, discordance was defined statistically from the joint distribution of apoB and LDL-C in this sample rather than by a clinical standard, and the threshold of an absolute residual *z* score of 1 is a convention.

## Conclusions

In a nationally representative sample of U.S. adults, the direction of apoB/LDL-C discordance was associated with systemic inflammation. Discordantly high apoB relative to LDL-C was associated with higher odds of elevated hs-CRP, while discordantly high LDL-C relative to apoB was associated with lower odds. These associations persisted after adjustment for age, sex, BMI, and diabetes, and the apoB-high association was stronger among statin-naive adults. Discordantly high apoB may therefore mark a phenotype in which excess atherogenic particle burden and low-grade inflammation coexist. Prospective studies are needed to determine whether this combination explains the excess cardiovascular risk linked to apoB-high discordance and whether it identifies patients who would benefit from both particle-lowering and anti-inflammatory therapy.

## Data Availability

All data referred to in this manuscript can be freely downloaded at https://wwwn.cdc.gov/nchs/nhanes/default.aspx

## Nonstandard Abbreviations and Acronyms

apoB: apolipoprotein B
ASCVD: atherosclerotic cardiovascular disease
hs-CRP: high-sensitivity C-reactive protein
LDL-C: low-density lipoprotein cholesterol
NHANES: National Health and Nutrition Examination Survey
OR: odds ratio

## Sources of Funding

None

## Disclosures

None

## Notes

### Competing Interest Statement

The authors have declared no competing interest.

### Author Declarations

This study is a secondary analysis of publicly available, de-identified data from NHANES 2015-2016. Because the data are public and contain no identifiable information, this analysis did not constitute human subjects research and did not require additional IRB review.

## References

[1] Scott M Grundy, Neil J Stone, Alison L Bailey, Craig Beam, Kim K Birtcher, Roger S Blumenthal, Lynne T Braun, Sarah De Ferranti, Joseph Faiella-Tommasino, Daniel E Forman, et al. 2018 aha/acc/aacvpr/aapa/abc/acpm/ada/ags/apha/aspc/nla/pcna guideline on the management of blood cholesterol: a report of the american college of cardiology/american heart association task force on clinical practice guidelines. circulation, 139(25):e1082–e1143, 2019.

[2] François Mach, Colin Baigent, Alberico L. Catapano, et al. 2019 ESC/EAS guidelines for the management of dyslipidaemias: lipid modification to reduce cardiovascular risk. European Heart Journal, 41:111–188, 2020.

[3] Writing Committee Members, Roger S Blumenthal, Pamela B Morris, Mario Gaudino, Heather M Johnson, Timothy S Anderson, Vera A Bittner, Ron Blankstein, LaPrincess C Brewer, Leslie Cho, et al. 2026 acc/aha/aacvpr/abc/acpm/ada/ags/apha/aspc/nla/pcna guideline on the management of dyslipidemia: a report of the american college of cardiology/american heart association joint committee on clinical practice guidelines. Circulation, 153(17):e1154–e1276, 2026.

[4] Brian A Ference, Henry N Ginsberg, Ian Graham, Kausik K Ray, Chris J Packard, Eric Bruckert, Robert A Hegele, Ronald M Krauss, Frederick J Raal, Heribert Schunkert, et al. Low-density lipoproteins cause atherosclerotic cardiovascular disease. 1. evidence from genetic, epidemiologic, and clinical studies. a consensus statement from the european atherosclerosis society consensus panel. European heart journal, 38(32):2459–2472, 2017.

[5] Allan D Sniderman, George Thanassoulis, Tamara Glavinovic, Ann Marie Navar, Michael Pencina, Alberico Catapano, and Brian A Ference. Apolipoprotein b particles and cardiovascular disease: a narrative review. JAMA cardiology, 4(12):1287–1295, 2019.

[6] John H Contois, Joseph P McConnell, Amar A Sethi, Gyorgy Csako, Sridevi Devaraj, Daniel M Hoefner, and G Russell Warnick. Apolipoprotein b and cardiovascular disease risk: position statement from the aacc lipoproteins and vascular diseases division working group on best practices, 2009.

[7] Allan D Sniderman, Ken Williams, John H Contois, Howard M Monroe, Matthew J McQueen, Jacqueline de Graaf, and Curt D Furberg. A meta-analysis of low-density lipoprotein cholesterol, non-high-density lipoprotein cholesterol, and apolipoprotein b as markers of cardiovascular risk. Circulation: Cardiovascular Quality and Outcomes, 4(3):337–345, 2011.

[8] John T. Wilkins, Rebecca C. Li, Allan Sniderman, Cheeling Chan, and Donald M. Lloyd-Jones. Discordance between apolipoprotein b and LDL-cholesterol in young adults predicts coronary artery calcification: the CARDIA study. Journal of the American College of Cardiology, 67: 193–201, 2016.

[9] Patrick R. Lawler, Akintunde O. Akinkuolie, Audrey Y. Chu, et al. Atherogenic lipoprotein determinants of cardiovascular disease and residual risk among individuals with low LDL cholesterol. Journal of the American Heart Association, 6:e005549, 2017.

[10] Allison W Peng, Eugenia Gianos, Michael D Shapiro, Ann Marie Navar, Sohail Zahid, Francoise A Marvel, Michael J Blaha, Fatima Rodriguez, Daniel E Soffer, Pamela B Morris, et al. Prevalence of apolipoprotein b and ldl cholesterol discordance: Insights from the vldbl and nhanes. Circulation, 153(4):290–293, 2026.

[11] Peter Libby. Inflammation in atherosclerosis. Arteriosclerosis, Thrombosis, and Vascular Biology, 32:2045–2051, 2012.

[12] Thomas A. Pearson, George A. Mensah, R. Wayne Alexander, et al. Markers of inflammation and cardiovascular disease: application to clinical and public health practice: a statement for healthcare professionals from the centers for disease control and prevention and the american heart association. Circulation, 107:499–511, 2003.

[13] Paul M. Ridker, Charles H. Hennekens, Julie E. Buring, and Nader Rifai. C-reactive protein and other markers of inflammation in the prediction of cardiovascular disease in women. New England Journal of Medicine, 342:836–843, 2000.

[14] Emerging Risk Factors Collaboration. C-reactive protein concentration and risk of coronary heart disease, stroke, and mortality: an individual participant meta-analysis. Lancet, 375: 132–140, 2010.

[15] Paul M. Ridker, Brendan M. Everett, Tom Thuren, et al. Antiinflammatory therapy with canakinumab for atherosclerotic disease. New England Journal of Medicine, 377:1119–1131, 2017.

[16] Jean-Claude Tardif, Simon Kouz, David D. Waters, et al. Efficacy and safety of low-dose colchicine after myocardial infarction. New England Journal of Medicine, 381:2497–2505, 2019.

[17] Stefan M. Nidorf, Aernoud T. L. Fiolet, Arend Mosterd, et al. Colchicine in patients with chronic coronary disease. New England Journal of Medicine, 383:1838–1847, 2020.

[18] Paul M. Ridker, Deepak L. Bhatt, Aruna D. Pradhan, et al. Inflammation and cholesterol as predictors of cardiovascular events among patients receiving statin therapy: a collaborative analysis of three randomised trials. Lancet, 401:1293–1301, 2023.

[19] George A Mensah, Natalie Arnold, Sumanth D Prabhu, Paul M Ridker, and Francine K Welty. Inflammation and cardiovascular disease: 2025 acc scientific statement: a report of the american college of cardiology. JACC, 2025.

[20] Anette Varbo, Marianne Benn, Anne Tybjærg-Hansen, and Børge G. Nordestgaard. Elevated remnant cholesterol causes both low-grade inflammation and ischemic heart disease, whereas elevated low-density lipoprotein cholesterol causes ischemic heart disease without inflammation. Circulation, 128:1298–1309, 2013.

[21] Børge G. Nordestgaard. Triglyceride-rich lipoproteins and atherosclerotic cardiovascular disease: new insights from epidemiology, genetics, and biology. Circulation Research, 118:547–563, 2016.

[22] Centers for Disease Control and Prevention, National Center for Health Statistics. National health and nutrition examination survey data. Hyattsville, MD: US Department of Health and Human Services. URL https://www.cdc.gov/nchs/nhanes/.

[23] Clifford L. Johnson, Ryne Paulose-Ram, Cynthia L. Ogden, et al. National health and nutrition examination survey: analytic guidelines, 1999–2010. Vital and Health Statistics, Series 2, (161): 1–24, 2013.

[24] Samia Mora, Julie E Buring, and Paul M Ridker. Discordance of low-density lipoprotein (ldl) cholesterol with alternative ldl-related measures and future coronary events. Circulation, 129 (5):553–561, 2014.

[25] Mohamed B Elshazly, Renato Quispe, Erin D Michos, Allan D Sniderman, Peter P Toth, Maciej Banach, Krishnaji R Kulkarni, Josef Coresh, Roger S Blumenthal, Steven R Jones, et al. Patient-level discordance in population percentiles of the total cholesterol to high-density lipoprotein cholesterol ratio in comparison with low-density lipoprotein cholesterol and non–high-density lipoprotein cholesterol: the very large database of lipids study (vldl-2b). Circulation, 132(8):667–676, 2015.

[26] Jing Cao, Sarah O Nomura, Brian T Steffen, Weihua Guan, Alan T Remaley, Amy B Karger, Pamela Ouyang, Erin D Michos, and Michael Y Tsai. Apolipoprotein b discordance with low-density lipoprotein cholesterol and non–high-density lipoprotein cholesterol in relation to coronary artery calcification in the multi-ethnic study of atherosclerosis (mesa). Journal of clinical lipidology, 14(1):109–121, 2020.

[27] Allan D Sniderman, Line Dufresne, Karol M Pencina, Selin Bilgic, George Thanassoulis, and Michael J Pencina. Discordance among apob, non–high-density lipoprotein cholesterol, and triglycerides: implications for cardiovascular prevention. European heart journal, 45(27): 2410–2418, 2024.

[28] Charles R Harper and Terry A Jacobson. Using apolipoprotein b to manage dyslipidemic patients: time for a change? In Mayo Clinic Proceedings, volume 85, pages 440–445. Elsevier, 2010.

[29] Nicholas A Marston, Robert P Giugliano, Giorgio EM Melloni, Jeong-Gun Park, Valerie Morrill, Michael A Blazing, Brian Ference, Evan Stein, Erik S Stroes, Eugene Braunwald, et al. Association of apolipoprotein b–containing lipoproteins and risk of myocardial infarction in individuals with and without atherosclerosis: distinguishing between particle concentration, type, and content. JAMA cardiology, 7(3):250–256, 2022.

[30] Amit Khera, Darren K. McGuire, Sabina A. Murphy, et al. Race and gender differences in c-reactive protein levels. Journal of the American College of Cardiology, 46:464–469, 2005.

[31] Susan G. Lakoski, Mary Cushman, Michael Criqui, et al. Gender and c-reactive protein: data from the multiethnic study of atherosclerosis (MESA) cohort. American Heart Journal, 152: 593–598, 2006.

